# *RIC3* variants are not associated with Parkinson’s Disease in large European, Latin American, or East Asian cohorts

**DOI:** 10.1101/2021.06.22.21259012

**Authors:** Kajsa Brolin, Sara Bandres-Ciga, Hampton Leonard, Mary B. Makarious, Cornelis Blauwendraat, Ignacio F Mata, Jia Nee Foo, Lasse Pihlstrøm, Maria Swanberg, Ziv Gan-Or, Manuela MX Tan, International Parkinson’s Disease Genomics Consortium (IPDGC)

**Affiliations:** Lund University, Translational Neurogenetics Unit, Wallenberg Neuroscience Center, Department of Experimental Medical Science, Lund, Sweden; Molecular Genetics Section, Laboratory of Neurogenetics, National Institute on Aging, National Institutes of Health, Bethesda, MD, 20892, USA; Center for Alzheimer’s and Related Dementias, National Institute on Aging, Bethesda, MD, 20892, USA; Data Tecnica International, Glen Echo, MD, 20812, USA; Department of Clinical and Movement Neurosciences, UCL Queen Square Institute of Neurology, University College London, London, UK; UCL Movement Disorders Centre, University College London, London, UK; Lerner Research Institute, Genomic Medicine, Cleveland Clinic Foundation, Cleveland, OH, USA; Lee Kong Chian School of Medicine, Nanyang Technological University Singapore, Singapore; Human Genetics, Genome Institute of Singapore, A*STAR, Singapore; Department of Neurology, Oslo University Hospital, Oslo, Norway; Department of Human Genetics, McGill University, Montreal, Quebec, Canada; Montreal Neurological Institute, McGill University, Montreal, Quebec, Canada; Department of Neurology and Neurosurgery, McGill University, Montreal, Quebec, Canada

**Author notes:** Corresponding authors: Kajsa Brolin,; Manuela MX Tan.

**Keywords:** Parkinson’s disease, *RIC3*, Genetics

## Abstract

Parkinson’s disease (PD) is a complex neurodegenerative disorder in which both rare and common genetic variants contribute to disease risk. Multiple genes have been reported to be linked to monogenic PD, but these only explain a fraction of the observed familial aggregation. Rare variants in *RIC3* have been suggested to be associated with PD in the Indian population. However, replication studies yielded inconsistent results. We further investigate the role of *RIC3* variants in PD in European cohorts using individual-level genotyping data from 14,671 PD patients and 17,667 controls, as well as whole-genome sequencing data from 1,615 patients and 961 controls. We also investigated *RIC3* using summary statistics from a Latin American cohort of 1,481 individuals, and from a cohort of 31,575 individuals of East Asian ancestry. We did not identify any association between *RIC3* and PD in any of the cohorts. However, more studies of rare variants in non-European ancestry populations, in particular South Asian populations, are necessary to further evaluate the world-wide role of *RIC3* in PD etiology.

## Introduction

Parkinson’s disease (PD) is a progressive neurodegenerative disorder, for which the genetic causes are largely still unknown. Variants in resistance to inhibitors of cholinesterase 3 (*RIC3)* were first identified in Indian PD patients (Sudhaman *et al*., 2016). A rare missense variant, p.P57T, was identified in a large Indian PD family with an autosomal-dominant pattern of inheritance, where it was present in all nine affected individuals and absent in five unaffected family members (Sudhaman *et al*., 2016). Another rare heterozygous missense *RIC3* variant, p.V168L, was found in the same study through targeted screening of an independent cohort of 288 Indian PD patients and 186 controls (Sudhaman *et al*., 2016). However, no association was found between *RIC3* variants and PD risk in later studies of French-Canadian and French cohorts (Ross *et al*., 2017) and a Chinese cohort (He *et al*., 2017). The *RIC3* gene encodes for a member of the resistance to inhibitors of cholinesterase 3-like family. It functions as a chaperone for nicotinic acetylcholine receptors, specifically the alpha-7 subunit of homomeric nicotinic acetylcholine receptors (CHRNA7). These receptors are important for promoting the release of dopamine in the nigrostriatal pathway (Gotti and Clementi, 2004; Quik and Kulak, 2002) where the degeneration and loss of dopaminergic neurons occurs in PD. In addition, there is some suggestion that smoking and nicotine are protective against PD (Breckenridge et al., 2016; Li et al., 2015). The original study showed that both variants in *RIC3* reduced the level of these receptors in mutant cell lines (Sudhaman *et al*., 2016). The authors suggest this is potentially through a dominant-negative effect of the mutations, as the level of CHRNA7 in the mutant cell lines was lower than in untransfected cells (Sudhaman *et al*., 2016).

Here, we assessed the role of *RIC3* variants in PD risk in several cohorts of different ethnicities: large European cohorts from the International Parkinson’s Disease Genomics Consortium (IPDGC) (Nalls et al., 2019) and the Accelerating Medicines Partnership in Parkinson’s Disease (AMP-PD) (https://amp-pd.org/). We further examined variants in *RIC3* in summary statistics from 1,481 individuals from the Latin American Research consortium on the Genetics of PD (LARGE-PD) (Loesch et al., 2020), as well as from 31,575 individuals of East Asian ancestry (Foo et al., 2020).

## Methods

We analyzed individual-level data from two large European datasets of PD patients and controls: IPDGC genome-wide genotyping data (14,671 PD patients and 17,667 controls), and AMP-PD whole-genome sequencing data (1,615 PD patients and 961 controls after quality control and removal of individuals of non-European ancestry [Supplementary Methods]). Analysis and quality control of the AMP-PD dataset (release 1) were performed on the cloud-native platform Terra (https://app.terra.bio/). In each dataset, GWAS analysis was performed using logistic regression in PLINK v1.9 (Chang et al., 2015).We adjusted for age at study entry, sex, and the first 10 genetic principal components (PCs). In both datasets, we also assessed the burden of rare variants in *RIC3*, using RVTESTS version 2.1.0 (Zhan et al., 2016) with standard settings. Variants were annotated using the latest available version (2019Oct24) of ANNOVAR (Wang et al., 2010). Only variants with minor allele frequency (MAF) less than 3% were included for burden analysis. All code for these analyses is available at: https://github.com/ipdgc/IPDGC-Trainees/blob/master/RIC3.ipynb. Subsequently, associations between variants in *RIC3* and PD were investigated in GWAS summary statistics from two large additional cohorts, LARGE-PD with 1,481 individuals (798 cases and 683 controls) (Loesch et al., 2020), and an Asian PD case-control cohort (Foo et al., 2020). The Asian PD GWAS included 31,575 individuals (6,724 PD patients and 24,851 controls) from East Asia, including Singapore/Malaysia, Hong Kong, Taiwan, mainland China, and South Korea (Foo et al., 2020). LocusZoom plots were generated using the LocalZoom tool available at https://my.locuszoom.org.

## Results

Using IPDGC genotyping data, we identified 143 variants within the *RIC3* gene (base pair coordinates 11:8,127,603-8,190,602 in genome build hg19/GRCh37; 11:8,106,056-8,169,055 in build GRCh38), including 3 missense variants (Supplementary Table 1). Neither of the two variants from the original study, p.P57T and p.V168L, were observed in this dataset. None of the *RIC3* variants were associated with PD risk after Bonferroni correction (p-value threshold 3.5 × 10^−4^, Figure 1A). We performed gene-based burden analysis of *RIC3* to assess the cumulative effect of rare variants (MAF < 3%). There was no significant association between *RIC3* and PD risk in burden analysis (N variants = 6, CMC p-value = 0.67, Fp p-value = 0.78, Madson-Browning p-value = 0.79, SKAT-O p-value = 0.79, Zeggini p-value = 0.76) (Supplementary Table 2). There were no missense variants in the genotyping data with MAF < 3%.

**Figure 1.**
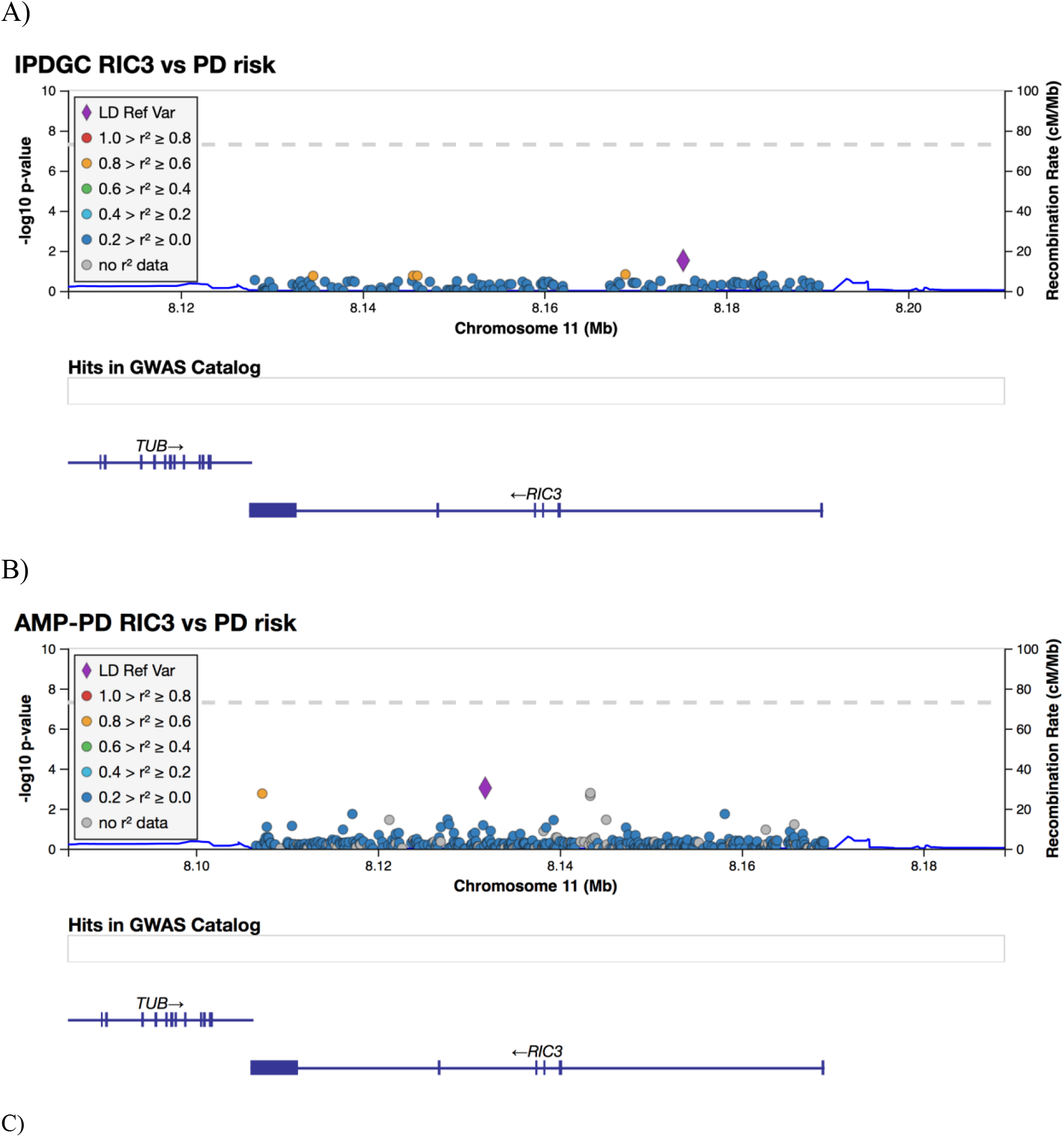
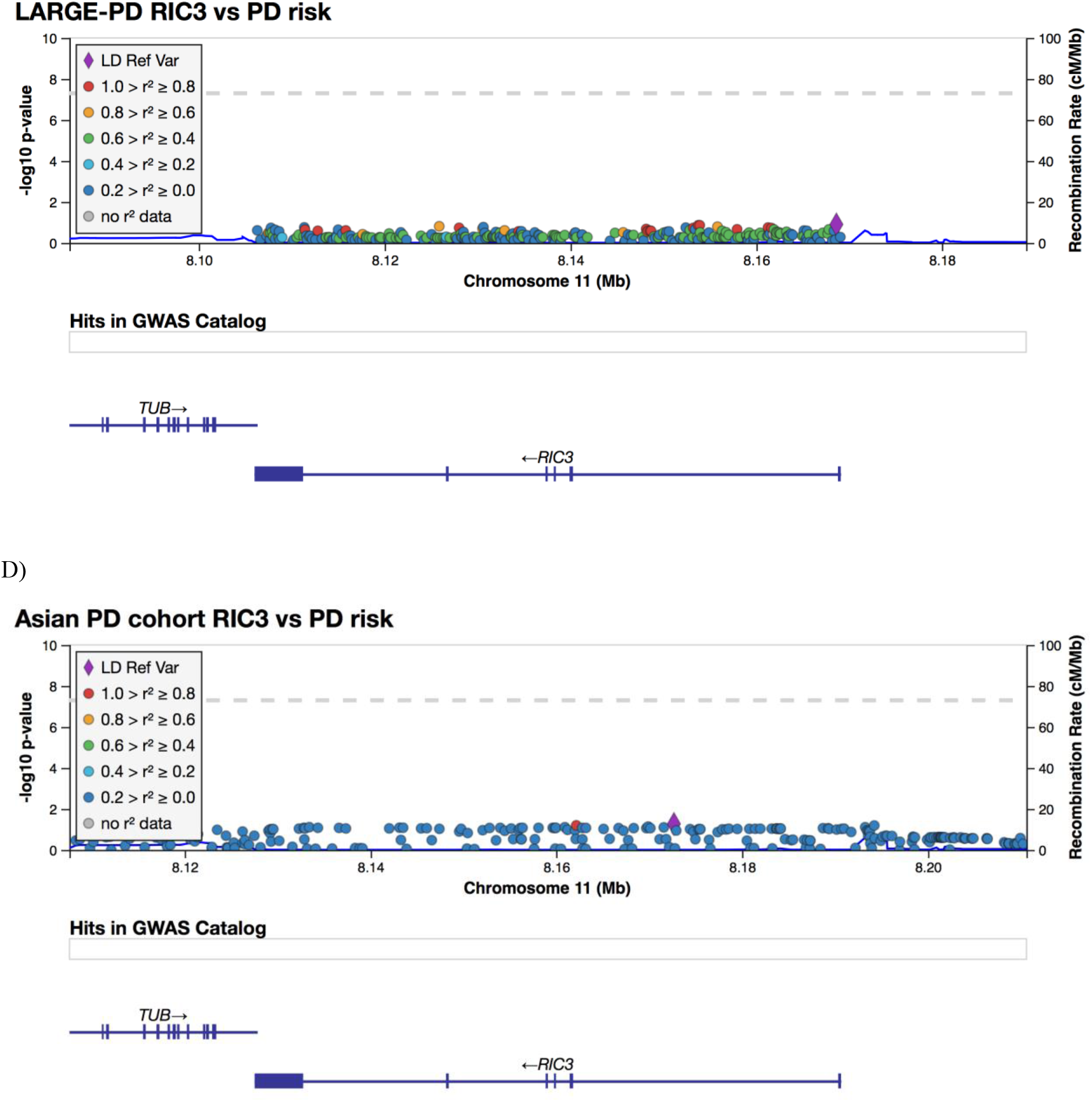
LocusZoom plots of the *RIC3* region, showing the p-values and recombination rate of the SNPs analyzed in the A) IPDGC genotyping data (hg19/GRCh37), B) AMP-PD WGS data (GRCh38), C) LARGE-PD GWAS summary statistic data (GRCh38), and D) East Asian GWAS summary statistic data (hg19/GRCh37). The variant with the lowest p-value from the logistic regression analyses is annotated with a purple diamond in each plot.

In the AMP-PD whole-genome sequencing data, we identified 773 variants in *RIC3*, including 14 missense variants and 5 synonymous variants (Supplementary Table 3). The two variants from the original study were not present in this dataset. No variants were associated with PD risk after Bonferroni correction (p-value threshold 5.1 × 10^−5^, Figure 1B).

There was no significant burden of rare variants in *RIC3* in association with PD risk using AMP-PD data (N variants = 571, all p-values > 0.05) (Supplementary Table 2). There was also no significant association between *RIC3* with PD risk when assessing rare coding variants only (N variants = 15, all p-values > 0.05).

In the LARGE-PD data, 293 variants with a MAF > 1% were identified in *RIC3* whereas in the Asian dataset, 126 variants were identified. None of the variants in *RIC3* in either of the datasets were associated with PD risk (Figure 1C and 1D).

## Discussion

We did not find any association between either single variants or the burden of rare variants in *RIC3* and PD risk in the European datasets. These are the largest publicly available European PD case-control cohorts that have been screened for *RIC3*. We also did not observe any associations between variants in *RIC3* and PD in GWAS summary statistics in cohorts from Latin America and East Asia. Overall, we did not find evidence to support the hypothesis that *RIC3* is associated with PD risk in individuals of European, Latin American, or East Asian ancestry.

However, we cannot rule out the possibility that rare *RIC3* variants may be associated with PD in specific families or populations. We did not identify the two originally reported variants p.P57T and p.V168L in WGS data, while the other datasets only included common variants. In addition, it is possible that only a few select rare variants in *RIC3* are pathogenic for PD, and this may not be detected by rare variant burden tests if the majority of other rare variants across the gene are not associated with PD.

It is also important to recognize that *RIC3* variants were identified in the Indian population as described in the original report, whereas our analysis of rare variants has been conducted in European populations. Only variants with a MAF > 1% were available in the GWAS summary statistics for the datasets of Latin American and Asian ancestry. It is possible that rare variants in this gene have a population-specific effect on PD risk. GWASs of PD in East Asian populations have identified novel variants for PD risk not found in European GWASs (Foo et al., 2020). In the Genome Aggregation Database (gnomAD) (Karczewski et al., 2020), both of the variants reported in the original study have higher allele frequencies in the South Asian population. For the p.P57T mutation (rs778138358), a total of 14 alleles were identified in 30,596 total alleles (MAF 0.05%). For the p.V168L mutation (rs777471396), 25 alleles were identified in the South Asian population out of a total of 30,614 alleles (MAF 0.08%). One allele carrier for each variant was also identified in the category group ‘Other’ (population not assigned). In contrast, the variants were absent in all other populations on gnomAD, including European, Latino, and East Asian. This suggests that the reported variants in *RIC3* are specific to South Asian populations, but does not answer the question of whether they are associated with PD risk, as individuals in gnomAD may not have been systematically screened for PD (Karczewski et al., 2020). Further studies or rare variants in South Asian populations are needed to clarify the role of *RIC3* variants in PD etiology.

In summary, we did not find evidence that *RIC3* variants are associated with PD risk in European cohorts. We also did not find evidence of more common variants (MAF > 1%) being associated with PD in cohorts from Latin America and East Asia. Based on the varying frequency of *RIC3* rare variants across geographic populations, further studies are encouraged of rare variants in *RIC3* in non-European populations, in particular South Asian populations, in order to fully evaluate the suggested role for *RIC3* in PD etiology.

## Data Availability

Data used in preparation of this article were obtained from the AMP-PD Knowledge Platform in which the clinical data and biosamples were obtained from the Fox Investigation for New Discovery of Biomarkers (BioFIND), the Harvard Biomarker Study (HBS), the Parkinson's Progression Markers Initiative (PPMI), and the Parkinson's Disease Biomarkers Program (PDBP). The investigators for each cohort have not participated in reviewing the data analysis or content of the manuscript.
All code for these analyses is available at: https://github.com/ipdgc/IPDGC-Trainees/blob/master/RIC3.ipynb.

https://www.amp-pd.org

## CRediT authorship contribution statement

**Kajsa Brolin:** Formal analysis, Software, Visualization, Writing - original draft. **Sara Bandres-Ciga:** Conceptualization, Formal analysis, Software, Data Curation, Supervision, Writing - review and editing. **Hampton Leonard:** Data curation, Software, Writing - review and editing. **Mary B. Makarious:** Data curation, Software, Writing - review and editing. **Cornelis Blauwendraat:** Investigation, Software, Data curation, Writing - review and editing. **Ignacio F Mata:** Investigation, Data curation, Writing - review and editing. **Jia Nee Foo:** Investigation, Data curation, Writing - review and editing. **Lasse Pihlstrøm:** Funding acquisition, Writing - review and editing. **Maria Swanberg:** Funding acquisition, Writing - review and editing. **Ziv Gan-Or:** Conceptualization, Writing - review and editing. **Manuela MX Tan:** Formal analysis, Software, Supervision, Writing - original draft.

## Acknowledgments

We would like to thank the participants who donated their time and biological samples, making this study possible. We would like to thank all members of the International Parkinson’s Disease Genomic Consortium (IPDGC), the Latin American Research consortium on the Genetics of PD (LARGE-PD), and the East Asian PD GWAS. For a complete overview of members, acknowledgments and funding, please see http://pdgenetics.org/partners and http://large-pd.org/. This work was supported in part by the Norwegian South-East Regional Health Authority (Helse Sør-Øst RHF) (supporting MMXT and LP) and in part by the Swedish Research Council (VR) and Åke Wiberg’s foundation (supporting KB and MS). We would also like to thank the Accelerating Medicines Partnership - Parkinson’s disease (AMP-PD). Data used in preparation of this article were obtained from the AMP-PD Knowledge Platform in which the clinical data and biosamples were obtained from the Fox Investigation for New Discovery of Biomarkers (BioFIND), the Harvard Biomarker Study (HBS), the Parkinson’s Progression Markers Initiative (PPMI), and the Parkinson’s Disease Biomarkers Program (PDBP). The investigators for each cohort have not participated in reviewing the data analysis or content of the manuscript. More information on the cohorts can be found in the supplementary material. Up-to-date information for AMP-PD can be found at https://www.amp-pd.org.

