## Supplementary material for "*RIC3* variants are not associated with Parkinson’s Disease in large European, Latin American, or East Asian cohorts"

### Introduction

Parkinson's disease (PD) is a complex and progressive neurodegenerative disorder where both rare and common genetic variants contribute to disease risk. Studying these genetic variants can provide important insights about the biological processes in PD pathology and etiology. Rare genetic variants with large effect sizes but with varying penetrance in several genes have been reported to be associated with monogenic PD. Although the understanding of the genetic basis of PD has increased significantly, replication and functional validation studies of novel genes associated with PD are still of great need and challenge (Bandres-Ciga et al., 2020; Blauwendraat et al., 2020).

*RIC3* is one gene reported to be associated with PD but where replication studies are needed. Variants in *RIC3* were first identified in Indian PD patients were a rare missense variant, p.P57T, was identified in a large autosomal dominant Indian PD family with nine affected individuals (Sudhaman et al., 2016). Another heterozygous missense *RIC3* variant, p.V168L, was found through targeted screening of an independent cohort of Indian PD patients (Sudhaman et al., 2016). However, no association was observed between *RIC3* variants and PD risk in later studies of French-Canadian and French cohorts (Ross et al., 2017) and a Chinese cohort (He et al., 2017).

The *RIC3* gene encodes the resistance to inhibitors of cholinesterase 3 protein. This acts as a chaperone for nicotinic acetylcholine receptors (nAChRs). There are many different types of nAChRs, and *RIC3* has been shown to have different effects on different types of nAChRs. Alternative splicing of *RIC3* could affect its functions, as *RIC3* isoforms have been shown to be differentially expressed in different tissues (Halevi et al., 2003). However, studies have shown that *RIC3* influences mainly the neuronal alpha-7 acetylcholine receptor (CHRNA7). This receptor is one of the primary nAChR subtypes in the brain (Quik et al., 2015). Many in-vitro studies have shown that *RIC3* affects CHRNA7 expression (Alexander et al., 2010; Ben-David and Treinin, 2017). In addition, the original report of *RIC3* in PD patients also showed that the two mutations observed in *RIC3* reduced the expression of CHRNA7 (Sudhaman et al., 2016).

nAChRs have been implicated in PD and other neurodegenerative disorders (Gotti and Clementi, 2004; Schaaf, 2014). The receptors are important for promoting the release of dopamine in the nigrostriatal pathway (Gotti and Clementi, 2004; Quik and Kulak, 2002) where the degeneration and loss of dopaminergic neurons occurs in PD. There is also consistent evidence of reduction of nicotine binding in the striatum of PD patients (Gotti and Clementi, 2004). In addition, studies have shown loss of nAChRs in the cerebral cortex (Burghaus et al., 2003), substantia nigra (Perry et al., 1995), and other brain regions in PD patients (Rinne et al., 1991). Finally, it has been suggested that nicotine and smoking have a protective effect against PD, with evidence from both animal studies (Quik and Kulak, 2002) and human epidemiological studies (Breckenridge et al., 2016; Li et al., 2015).

Thus, it is plausible that *RIC3* is involved in PD pathogenesis through the alteration of nAChRs, in particular the CHRNA7 subtype (Quik et al., 2015). However, so far only one study has reported an association between *RIC3* mutations and PD in an Indian population (Sudhaman et al., 2016). Additional studies have not confirmed this association in other populations, including in French-Canadians (Ross et al., 2017) and Chinese populations (He et al., 2017). Therefore, further analysis of large cohorts is needed to determine whether *RIC3* mutations are associated with PD risk. We aimed to investigate whether *RIC3* variants are associated with PD using data from large European cohorts. In addition, we investigated associations of variants in *RIC3* and PD using GWAS summary statistics from two additional cohorts of Latin American (Loesch et al., 2020) and East Asian ancestry (Foo et al., 2020).

### Methods

Genotyping data from 4,671 PD patients and 17,667 control from the IPDGC was used for the analyses. Quality control (QC), imputation and post-imputation QC had been performed prior to the analyses as previously described (Nalls et al., 2019). In addition, publicly available whole-genome sequencing (WGS) data from the Accelerating Medicines Partnership - Parkinson's Disease (AMP-PD) was accessed through and analyzed on the cloud-native platform Terra provided by Broad institute (https://app.terra.bio/). The available data consisted of a total of 1,735 individuals with a PD diagnosis and 1,100 individuals without a PD diagnosis (controls). More information on the AMP-PD WGS data can be found at: https://amp-pd.org/whole-genome-data.

We performed additional QC of the AMP-PD WGS data using PLINK 1.9 (Chang et al., 2015; Purcell et al., 2007). Sample QC included exclusion of samples with a genotyping call rate < 95%, exclusion of samples exhibiting excessive heterozygosity estimated by a F statistic of < -0.15 or > 0.15 and exclusion of samples whose genetically determined sex did not match the sex reported in the clinical data. Sex was determined genetically using X-chromosome heterogeneity analysis where a male call was reported if the X-chromosome inbreeding (homozygosity) estimate (F) was > 0.75 and a female call if F < 0.25. Ancestry outliers and closely related samples were also excluded from the analyses. Ancestry outliers were identified using principal component analysis (PCA) with the HapMap phase 3 data as a reference dataset (International HapMap Consortium, 2003) (https://www.sanger.ac.uk/resources/downloads/human/hapmap3.html). Samples clustering 6 standard deviations (SD +/-) around the combined population mean of the European ancestry populations Utah residents with Northern and Western European ancestry (CEU) and Toscani in Italia (TSI) were kept for further analyses. Samples closely related, defined as sharing > 12.5% of alleles were also excluded. Following sample QC, variant QC was performed where variants with a missingness rate > 5% or with a Hardy-Weinberg equilibrium p-value < 0.0001 were excluded. Differences in missingness for variants was also investigated between PD patents and controls and by haplotypes where variants with a p-value <= 0.0001 were excluded. A total of 1,615 patients and 961 controls passed the quality control (QC). Mean age at baseline for these study participants (defined as the difference between date of birth (years) and informed consent date) was 64.3 (SD±9.5) years for the patient group and 60.9 (SD±11.6) years for the control group. A subsequent PCA was performed in order to generate principal components to use in following regression analyses to adjust for population stratification.

The region for *RIC3* was extracted from both the genotyping and WGS datasets using the Ensembl gene coordinates (11:8,127,603-8,190,602 in genome build hg19/GRCh37; 11:8,106,056-8,169,055 in build GRCh38). The region was annotated using ANNOVAR (Wang et al., 2010). Logistic regression was used to test for associations between *RIC3* variants and PD risk with adjustment for sex, age, and genetic PC1-PC10. Variants in *RIC3* with a minor allele frequency (MAF) of ≤ 3% were used in gene-based burden analysis tests in order to assess the cumulative effect of multiple low frequency variants. The burden analyses were performed in RVTESTS version 2.1.0 (Zhan et al., 2016) with both datasets and included the sequence Kernel association test (SKAT), optimized SKAT (SKAT-O), CMC, Zeggini,

Madson-Browning, and Fp. These are different methods of aggregating rare variants. Additionally, the SKAT methods are suitable to test rare variants with different directions of effects (Zhan et al., 2016). All code can be found at https://github.com/ipdgc/IPDGC-Trainees/blob/master/RIC3.ipynb

Moreover, we looked at variants in *RIC3* in GWAS summary statistics in data from two additional cohorts of Latin American and Asian ancestry. These cohorts comprised of data from 1,481 individuals (798 cases and 683 controls) from the Latin American Research consortium on the Genetics of PD (LARGE-PD) (Loesch et al., 2020), and from 31,575 individuals (6,724 PD patients and 24,851 controls) of Asian ancestry (Foo et al., 2020). LocusZoom plots were generated for all cohorts using the LocalZoom tool available at https://my.locuszoom.org.

**Supplementary Acknowledgments**

We would like to thank the Accelerating Medicines Partnership - Parkinson's disease (AMP-PD), a public-private partnership between the National Institute of Neurological Disorders and Stroke (NINDS), the Food and Drug Administration (FDA), GSK, Pfizer, Sanofi, Celgene, Verily and the Michael J. Fox Foundation (MJFF) managed through the Foundation for the NIH (FNIH). Clinical data and biosamples used in preparation of this article were obtained from the Fox Investigation for New Discovery of Biomarkers (BioFIND), the Harvard Biomarker Study (HBS), the Parkinson's Progression Markers Initiative (PPMI), and the Parkinson's Disease Biomarkers Program (PDBP). BioFIND is sponsored by The Michael J. Fox Foundation for Parkinson's Research (MJFF) with support from the National Institute for Neurological Disorders and Stroke (NINDS). The BioFIND Investigators have not participated in reviewing the data analysis or content of the manuscript. For up-to-date information on the study, visit michaeljfox.org/biofind. Harvard Biomarker Study (HBS) is a collaboration of HBS investigators [full list of HBS investigators found at https://www.bwhparkinsoncenter.org/biobank] and funded through philanthropy and NIH and Non-NIH funding sources. The HBS Investigators have not participated in reviewing the data analysis or content of the manuscript. PPMI – a public-private partnership – is funded by the Michael J. Fox Foundation for Parkinson's Research and funding partners, (www.ppmi-info.org/fundingpartners). The PPMI Investigators have not participated in reviewing the data analysis or content of the manuscript. For up-to-date information on the study, visit www.ppmi-info.org. Parkinson's Disease Biomarker Program (PDBP) consortium is supported by the National Institute of Neurological Disorders and Stroke (NINDS) at the National Institutes of Health. A full list of PDBP investigators can be found at https://pdbp.ninds.nih.gov/policy. The PDBP Investigators have not participated in reviewing the data analysis or content of the manuscript. This research was supported, in part, by the Intramural Research Program of the National Institutes of Health (National Institute on Aging) project numbers 1ZIA- NS003154, Z01- AG000949- 02, and Z01- ES10198.

Karczewski, K.J., Francioli, L.C., Tiao, G., Cummings, B.B., Alföldi, J., Wang, Q., Collins, R.L., Laricchia, K.M., Ganna, A., Birnbaum, D.P., Gauthier, L.D., Brand, H., Solomonson, M., Watts, N.A., Rhodes, D., Singer-Berk, M., England, E.M., Seaby, E.G., Kosmicki, J.A., Walters, R.K., Tashman, K., Farjoun, Y., Banks, E., Poterba, T., Wang, A., Seed, C., Whiffin, N., Chong, J.X., Samocha, K.E., Pierce-Hoffman, E., Zappala, Z., O’Donnell-Luria, A.H., Minikel, E.V., Weisburd, B., Lek, M., Ware, J.S., Vittal, C., Armean, I.M., Bergelson, L., Cibulskis, K., Connolly, K.M., Covarrubias, M., Donnelly, S., Ferriera, S., Gabriel, S., Gentry, J., Gupta, N., Jeandet, T., Kaplan, D., Llanwarne, C., Munshi, R., Novod, S., Petrillo, N., Roazen, D., Ruano-Rubio, V., Saltzman, A., Schleicher, M., Soto, J., Tibbetts, K., Tolonen, C., Wade, G., Talkowski, M.E., Aguilar Salinas, C.A., Ahmad, T., Albert, C.M., Ardissino, D., Atzmon, G., Barnard, J., Beaugerie, L., Benjamin, E.J., Boehnke, M., Bonnycastle, L.L., Bottinger, E.P., Bowden, D.W., Bown, M.J., Chambers, J.C., Chan, J.C., Chasman, D., Cho, J., Chung, M.K., Cohen, B., Correa, A., Dabelea, D., Daly, M.J., Darbar, D., Duggirala, R.,

Dupuis, J., Ellinor, P.T., Elosua, R., Erdmann, J., Esko, T., Färkkilä, M., Florez, J., Franke, A., Getz, G., Glaser, B., Glatt, S.J., Goldstein, D., Gonzalez, C., Groop, L., Haiman, C., Hanis, C., Harms, M., Hiltunen, M., Holi, M.M., Hultman, C.M., Kallela, M., Kaprio, J., Kathiresan, S., Kim, B.J., Kim, Y.J., Kirov, G., Kooner, J., Koskinen, S., Krumholz, H.M., Kugathasan, S., Kwak, S.H., Laakso, M., Lehtimäki, T., Loos, R.J.F., Lubitz, S.A., Ma, R.C.W., MacArthur, D.G., Marrugat, J., Mattila, K.M., McCarroll, S., McCarthy, M.I., McGovern, D., McPherson, R., Meigs, J.B., Melander, O., Metspalu, A., Neale, B.M., Nilsson, P.M., O'Donovan, M.C., Ongur, D., Orozco, L., Owen, M.J., Palmer, C.N.A., Palotie, A., Park, K.S., Pato, C., Pulver, A.E., Rahman, N., Remes, A.M., Rioux, J.D., Ripatti, S., Roden, D.M., Saleheen, D., Salomaa, V., Samani, N.J., Scharf, J., Schunkert, H., Shoemaker, M.B., Sklar, P., Soininen, H., Sokol, H., Spector, T., Sullivan, P.F., Suvisaari, J., Tai, E.S., Teo, Y.Y., Tiinamaija, T., Tsuang, M., Turner, D., Tusie-Luna, T., Vartiainen, E., Ware, J.S., Watkins, H., Weersma, R.K., Wessman, M., Wilson, J.G., Xavier, R.J., Neale, B.M., Daly, M.J., MacArthur, D.G., 2020. The mutational constraint spectrum quantified from variation in 141,456 humans. Nature 581, 434–443. https://doi.org/10.1038/s41586-020-2308-7

Li, X., Li, W., Liu, G., Shen, X., Tang, Y., 2015. Association between cigarette smoking and Parkinson's disease: A meta-analysis. Arch. Gerontol. Geriatr. 61, 510–516. https://doi.org/10.1016/j.archger.2015.08.004

Loesch, D., Horimoto, A.R.V.R., Heilbron, K., Sarihan, E.I., Inca-Martinez, M., Mason, E., Cornejo-Olivas, M., Torres, L., Mazzetti, P., Cosentino, C., Sarapura-Castro, E., Rivera-Valdivia, A., Medina, A.C., Dieguez, E., Raggio, V., Lescano, A., Tumas, V., Borges, V., Ferraz, H.B., Rieder, C.R., Schumacher-Schuh, A., Santos-Lobato, B.L., Velez-Pardo, C., Jimenez-Del-Rio, M., Lopera, F., Moreno, S., Chana-Cuevas, P., Fernandez, W., Arboleda, G., Arboleda, H., Arboleda-Bustos, C.E., Yearout, D., Zabetian, C.P., The 23andMe Research Team, Cannon, P., Thornton, T.A., O'Connor, T.D., Mata, I.F., on behalf of the Latin American Research Consortium on the Genetics of Parkinson's Disease (LARGE-PD), 2020. Characterizing the genetic architecture of Parkinson's disease in Latinos. medRxiv November.

Nalls, M.A., Blauwendraat, C., Vallerga, C.L., Heilbron, K., Bandres-Ciga, S., Chang, D., Tan, M., Kia, D.A., Noyce, A.J., Xue, A., Bras, J., Young, E., von Coelln, R., Simón-Sánchez, J., Schulte, C., Sharma, M., Krohn, L., Pihlstrøm, L., Siitonen, A., Iwaki, H., Leonard, H., Faghri, F., Gibbs, J.R., Hernandez, D.G., Scholz, S.W., Botia, J.A., Martinez, M., Corvol, J.C., Lesage, S., Jankovic, J., Shulman, L.M., Sutherland, M.,

Tienari, P., Majamaa, K., Toft, M., Andreassen, O.A., Bangale, T., Brice, A., Yang, J., Gan-Or, Z., Gasser, T., Heutink, P., Shulman, J.M., Wood, N.W., Hinds, D.A., Hardy, J.A., Morris, H.R., Gratten, J., Visscher, P.M., Graham, R.R., Singleton, A.B., Adarmes-Gómez, A.D., Aguilar, M., Aitkulova, A., Akhmetzhanov, V., Alcalay, R.N., Alvarez, I., Alvarez, V., Barrero, F.J., Bergareche Yarza, J.A., Bernal-Bernal, I., Billingsley, K., Blazquez, M., Bonilla-Toribio, M., Botía, J.A., Boungiorno, M.T., Brockmann, K., Bubb, V., Buiza-Rueda, D., Cámara, A., Carrillo, F., Carrión-Claro, M., Cerdan, D., Chelban, V., Clarimón, J., Clarke, C., Compta, Y., Cookson, M.R., Craig, D.W., Danjou, F., Diez-Fairen, M., Dols-Icardo, O., Duarte, J., Duran, R., Escamilla-Sevilla, F., Escott-Price, V., Ezquerra, M., Feliz, C., Fernández, M., Fernández-Santiago, R., Finkbeiner, S., Foltynie, T., Garcia, C., García-Ruiz, P., Gomez Heredia, M.J., Gómez-Garre, P., González, M.M., Gonzalez-Aramburu, I., Guelfi, S., Guerreiro, R., Hardy, J., Hassin-Baer, S., Hoenicka, J., Holmans, P., Houlden, H., Infante, J., Jesús, S., Jimenez-Escrig, A., Kaishybayeva, G., Kaiyrzhanov, R., Karimova, A., Kinghorn, K.J., Koks, S., Kulisevsky, J., Labrador-Espinosa, M.A., Leonard, H.L., Lewis, P., Lopez-Sendon, J.L., Lovering, R., Lubbe, S., Lungu, C., Macias, D., Manzoni, C., Marín, J., Marinus, J., Marti, M.J., Martínez Torres, I., Martínez-Castrillo, J.C., Mata, M., Mencacci, N.E., Méndez-del-Barrio, C., Middlehurst, B., Mínguez, A., Mir, P., Mok, K.Y., Muñoz, E., Narendra, D., Ojo, O.O., Okubadejo, N.U., Pagola, A.G., Pastor, P., Perez Errazquin, F., Periñán-Tocino, T., Pihlstrom, L., Plun-Favreau, H., Quinn, J., R'Bibo, L., Reed, X., Rezola, E.M., Rizig, M., Rizzu, P., Robak, L., Rodriguez, A.S., Rouleau, G.A., Ruiz-Martínez, J., Ruz, C., Ryten, M., Sadykova, D., Schreglmann, S., Shashkin, C., Sierra, M., Suarez-Sanmartin, E., Taba, P., Tabernero, C., Tan, M.X., Tartari, J.P., Tejera-Parrado, C., Tolosa, E., Trabzuni, D., Valldeoriola, F., van Hilten, J.J., Van Keuren-Jensen, K., Vargas-González, L., Vela, L., Vives, F., Williams, N., Zharkinbekova, N., Zharmukhanov, Z., Zholdybayeva, E., Zimprich, A., Ylikotila, P., Shulman, L.M., Reich, S., Savitt, J., Agee, M., Alipanahi, B., Auton, A., Bell, R.K., Bryc, K., Elson, S.L., Fontanillas, P., Furlotte, N.A., Huber, K.E., Hicks, B., Jewett, E.M., Jiang, Y., Kleinman, A., Lin, K.H., Litterman, N.K., McCreight, J.C., McIntyre, M.H., McManus, K.F., Mountain, J.L., Noblin, E.S., Northover, C.A.M., Pitts, S.J., Poznik, G.D., Sathirapongsasuti, J.F., Shelton, J.F., Shringarpure, S., Tian, C., Tung, J., Vacic, V., Wang, X., Wilson, C.H., Anderson, T., Bentley, S., Dalrymple-Alford, J., Fowdar, J., Halliday, G., Henders, A.K., Hickie, I., Kassam, I., Kennedy, M., Kwok, J., Lewis, S., Mellick, G., Montgomery, G., Pearson, J., Pitcher, T., Sidorenko, J., Silburn, P.A.,

Vallerga, C.L., Visscher, P.M., Wallace, L., Wray, N.R., Zhang, F., 2019. Identification of novel risk loci, causal insights, and heritable risk for Parkinson's disease: a meta-analysis of genome-wide association studies. Lancet Neurol. 18, 1091–1102. https://doi.org/10.1016/S1474-4422(19)30320-5

Perry, E.K., Morris, C.M., Court, J.A., Cheng, A., Fairbairn, A.F., McKeith, I.G., Irving, D., Brown, A., Perry, R.H., 1995. Alteration in nicotine binding sites in Parkinson's disease, Lewy body dementia and Alzheimer's disease: Possible index of early neuropathology. Neuroscience 64, 385–395. https://doi.org/10.1016/0306-4522(94)00410-7

Purcell, S., Neale, B., Todd-Brown, K., Thomas, L., Ferreira, M.A.R., Bender, D., Maller, J., Sklar, P., De Bakker, P.I.W., Daly, M.J., Sham, P.C., 2007. PLINK: A tool set for whole-genome association and population-based linkage analyses. Am. J. Hum. Genet. 81, 559–575. https://doi.org/10.1086/519795

Quik, M., Kulak, J.M., 2002. Nicotine and nicotinic receptors; relevance to Parkinson's disease. Neurotoxicology 23, 581–594. https://doi.org/10.1016/S0161-813X(02)00036-0

Quik, M., Zhang, D., McGregor, M., Bordia, T., 2015. Alpha7 nicotinic receptors as therapeutic targets for Parkinson's disease. Biochem. Pharmacol. 97, 399–407. https://doi.org/10.1016/j.bcp.2015.06.014

Rinne, J.O., Myllykylä, T., Lönnberg, P., Marjamäki, P., 1991. A postmortem study of brain nicotinic receptors in Parkinson's and Alzheimer's disease. Brain Res. 547, 167–170. https://doi.org/10.1016/0006-8993(91)90588-M

Ross, J.P., Dupré, N., Dauvilliers, Y., Strong, S., Dionne-Laporte, A., Dion, P.A., Rouleau, G.A., Gan-Or, Z., 2017. RIC3 variants are not associated with Parkinson's disease in French-Canadians and French. Neurobiol. Aging 53, 194.e9-194.e11. https://doi.org/10.1016/j.neurobiolaging.2017.01.005

Schaaf, C.P., 2014. Nicotinic acetylcholine receptors in human genetic disease. Genet. Med. 16, 649–656. https://doi.org/10.1038/gim.2014.9

Sudhaman, S., Muthane, U.B., Behari, M., Govindappa, S.T., Juyal, R.C., Thelma, B.K., 2016. Evidence of mutations in RIC3 acetylcholine receptor chaperone as a novel cause of autosomal-dominant Parkinson's disease with non-motor phenotypes. J. Med. Genet. 53, 559–566. https://doi.org/10.1136/jmedgenet-2015-103616

Wang, K., Li, M., Hakonarson, H., 2010. ANNOVAR : functional annotation of genetic variants from high-throughput sequencing data. Nucleic Acids Res. 38, e164. https://doi.org/10.1093/nar/gkq603

Zhan, X., Hu, Y., Li, B., Abecasis, G.R., Liu, D.J., 2016. RVTESTS: An efficient and comprehensive tool for rare variant association analysis using sequence data. Bioinformatics 32, 1423–1426. https://doi.org/10.1093/bioinformatics/btw079

**Supplementary Tables**

**Supplementary Table 1: Coding variants in IPDGC genotyping data**

**Supplementary Table 2: Burden analysis results**

**Supplementary Table 3: Coding variants in AMP-PD whole genome sequencing data**

Supplementary table 1

**Supplementary Table 1. Exonic variants in RIC3 from IPDGC data, with results from the logistic regression GWAS and allele frequencies.**

| Chr | BP | SNP | A1 | A2 | OR | STAT | P | Variant_Type | Gene | Consequence | Frequency_gnomAD_NFE | MAF_affected | MAF_unaffected | Amino acid change (canonical transcript) |
|---|---|---|---|---|---|---|---|---|---|---|---|---|---|---|
| 11 | 8132301 | rs11826236 | T | C | 1.029 | 0.734 | 0.463 | exonic | RIC3 | nonsynonymous SNV | 0.066 | 0.059 | 0.059 | RIC3:NM_001206671:exon6:c.G1054A:p.D352N |
| 11 | 8159843 | rs73411617 | A | G | 0.997 | -0.052 | 0.958 | exonic | RIC3 | nonsynonymous SNV | 0.033 | 0.034 | 0.033 | RIC3:NM_001206671:exon3:c.C403T:p.P135S |
| 11 | 8159857 | rs55990541 | T | C | 1.031 | 0.775 | 0.438 | exonic | RIC3 | nonsynonymous SNV | 0.066 | 0.059 | 0.059 | RIC3:NM_001206671:exon3:c.G389A:p.C130Y |
| 11 | 8159892 | rs10839976 | T | G | 0.979 | -0.955 | 0.34 | exonic | RIC3 | synonymous SNV | 0.242 | 0.224 | 0.236 | RIC3:NM_001206671:exon3:c.C354A:p.L118L |

Key: IPDGC, International Parkinson's Disease Genomics Consortium; MAF, Minor Allele Frequency; NFE, Non-Finnish Europeans in gnomAD; SNV, Single Nucleotide Variant

Supplementary table 2

| Supplementary Table 2. Rare variant burden test p-values for all variants and coding variants. Minor allele frequency threshold was | | | | | | | |
|---|---|---|---|---|---|---|---|
| | No. variants | CMC | Fp | Madson-Browni | SKAT | SKAT-O | Zeggini |
| **IPDGC** | | | | | | | |
| All variants | 6 | 0.674 | 0.784 | 0.789 | 0.794 | 1 | 0.756 |
| Coding variants c | 0 | NA | NA | NA | NA | NA | NA |
| **AMP-PD** | | | | | | | |
| All variants | 571 | 0.766 | 0.603 | 0.561 | 0.881 | 0.766 | 0.694 |
| Coding variants c | 15 | 0.291 | 0.081 | 0.51 | 0.624 | 0.568 | 0.334 |
| Key: SKAT, Sequence Kernel Association Test; SKAT-O, Optimized SKAT. | | | | | | | |

Supplementary table 3

**Supplementary Table 3. Exonic variants in RIC3 from AMP-PD WGS data, with results from the logistic regression analysis and allele frequencies.**

| Chr | BP | SNP | A1 | A2 | OR | SE | P | Variant_Type | Gene | Consequence | Frequency_gnomAD_NFE | MAF_all | MAF_affected | MAF_unaffected | CADD_phred | Amino acid change (canonical transcript) |
|---|---|---|---|---|---|---|---|---|---|---|---|---|---|---|---|---|
| 11 | 8110754 | rs11826236 | T | C | 1.039 | 0.139 | 0.783 | exonic | RIC3 | nonsynonymous SNV | 0.066 | 0.056 | 0.056 | 0.056 | 4.54 | RIC3:NM_001206671:exon6:c.G1054A:p.D352N |
| 11 | 8110755 | rs17852022 | A | G | 0.517 | 1.188 | 0.578 | exonic | RIC3 | synonymous SNV | 0.000 | 0.001 | 0 | 0.002 | . | RIC3:NM_001206671:exon6:c.C1053T:p.T351T |
| 11 | 8110798 | rs372177118 | G | T | NA | NA | NA | exonic | RIC3 | nonsynonymous SNV | 0.000 | 0 | 0 | 0.001 | 0.17 | RIC3:NM_001206671:exon6:c.A1010C:p.E337A |
| 11 | 8110874 | rs116932252 | T | C | 1.096 | 0.458 | 0.841 | exonic | RIC3 | nonsynonymous SNV | 0.008 | 0.005 | 0.005 | 0.005 | 18.44 | RIC3:NM_001206671:exon6:c.G934A:p.D312N |
| 11 | 8111075 | rs369921419 | C | T | NA | NA | NA | exonic | RIC3 | nonsynonymous SNV | 0.000 | 0 | 0 | 0.000 | 19.94 | RIC3:NM_001206671:exon6:c.A733G:p.T245A |
| 11 | 8126740 | rs139685245 | A | C | 0.321 | 1.041 | 0.276 | exonic | RIC3 | nonsynonymous SNV | 0.004 | 0.001 | 0.001 | 0.001 | 24.9 | RIC3:NM_001206671:exon5:c.G589T:p.V197F |
| 11 | 8126745 | rs150042716 | C | G | NA | NA | NA | exonic | RIC3 | nonsynonymous SNV | . | 0 | 0 | 0.000 | 22.5 | RIC3:NM_001206671:exon5:c.C584G:p.T195S |
| 11 | 8126754 | rs144870134 | T | C | NA | NA | NA | exonic | RIC3 | nonsynonymous SNV | 0.000 | 0 | 0 | 0.000 | 16.16 | RIC3:NM_001206671:exon5:c.G575A:p.R192Q |
| 11 | 8126885 | rs140788261 | G | C | NA | NA | NA | exonic | RIC3 | nonsynonymous SNV | 0.000 | 0 | 0 | 0.000 | 10.74 | *RIC3:NM_001346693:exon6:c.G381C:p.Q127H |
| 11 | 8137395 | rs201157012 | A | C | NA | NA | NA | exonic | RIC3 | synonymous SNV | 0.000 | 0 | 0 | 0.000 | . | RIC3:NM_001206671:exon4:c.G504T:p.V168V |
| 11 | 8138296 | rs73411617 | A | G | 0.978 | 0.151 | 0.885 | exonic | RIC3 | nonsynonymous SNV | 0.033 | 0.046 | 0.042 | 0.052 | 9.65 | RIC3:NM_001206671:exon3:c.C403T:p.P135S |
| 11 | 8138310 | rs55990541 | T | C | 1.011 | 0.136 | 0.935 | exonic | RIC3 | nonsynonymous SNV | 0.066 | 0.057 | 0.056 | 0.058 | 1.25 | RIC3:NM_001206671:exon3:c.G389A:p.C130Y |
| 11 | 8138337 | rs80168649 | G | C | 0.88 | 0.342 | 0.709 | exonic | RIC3 | nonsynonymous SNV | 0.014 | 0.009 | 0.009 | 0.009 | 21.8 | RIC3:NM_001206671:exon3:c.G362C:p.G121A |
| 11 | 8138345 | rs10839976 | T | G | 0.953 | 0.076 | 0.528 | exonic | RIC3 | synonymous SNV | 0.242 | 0.229 | 0.228 | 0.229 | . | RIC3:NM_001206671:exon3:c.C354A:p.L118L |
| 11 | 8140017 | rs144806410 | A | G | 0.704 | 0.441 | 0.426 | exonic | RIC3 | nonsynonymous SNV | 0.004 | 0.005 | 0.004 | 0.006 | 26.3 | RIC3:NM_001206671:exon2:c.C301T:p.P101S |
| 11 | 8140021 | rs201908482 | T | A | NA | NA | NA | exonic | RIC3 | synonymous SNV | 0.000 | 0 | 0 | 0.001 | . | RIC3:NM_001206671:exon2:c.T297A:p.I99I |
| 11 | 8140099 | rs149313414 | C | G | NA | NA | NA | exonic | RIC3 | synonymous SNV | 0.000 | 0 | 0 | 0.000 | . | RIC3:NM_001206671:exon2:c.C219G:p.A73A |
| 11 | 8168901 | rs202021657 | T | C | NA | NA | NA | exonic | RIC3 | nonsynonymous SNV | 0.000 | 0 | 0 | 0.001 | 15.27 | RIC3:NM_001206671:exon1:c.G89A:p.R30H |
| 11 | 8168916 | rs145965152 | C | T | 2.231 | 1.12 | 0.474 | exonic | RIC3 | nonsynonymous SNV | 0.002 | 0.001 | 0.002 | 0.001 | 14.63 | RIC3:NM_001206671:exon1:c.A74G:p.K25R |

Key: CADD, Combined Annotation Dependent Depletion; MAF, Minor Allele Frequency; NFE, Non-Finnish Europeans in gnomAD; SNV, Single Nucleotide Variant

*This variant only resulted in an amino acid change in a non-canonical transcript
